# Design tensions in a two-sided marketplace for reusable digital therapeutics software components: a qualitative interview study

**DOI:** 10.64898/2026.07.17.26358332

**Authors:** Sabra Melamed, Yannick Merz, Marcia Nißen, Tobias Kowatsch

**Affiliations:** Institute for Implementation Science in Health Care, University of Zurich, Zurich, Switzerland; School of Medicine, University of St. Gallen, St. Gallen, Switzerland; Centre for Digital Health Interventions, Department of Management, Technology, and Economics, ETH Zurich, Zurich, Switzerland; School of Health Sciences, Eastern Switzerland University of Applied Sciences (OST), St. Gallen, Switzerland

**Author notes:** **Corresponding author:** Prof. Dr. Tobias Kowatsch, Institute for Implementation Science in Health Care, University of Zurich, Zurich, Switzerland.

**Keywords:** Digital Health, Therapeutics, Software Design, Qualitative Research, Health Information Exchange

## Abstract

**Objectives:** To identify stakeholder-perceived design tensions in a two-sided marketplace for reusable digital therapeutics (DTx) software components and to use these tensions to propose alternative marketplace concepts.

**Methods:** We conducted 24 semi-structured interviews with digital health researchers and professionals. Data were analysed using hybrid deductive-inductive codebook thematic analysis. The Magic Triangle provided the initial deductive structure. One researcher coded all transcripts; a second independently applied the developing codebook to five transcripts to refine definitions and consistency. Seventeen parent themes were synthesized into 12 design tensions, which informed three author-generated marketplace concepts.

**Results:** Participants described trade-offs concerning target users and host, component scope and customization, quality labels, verification, geographic scope, pricing, interoperability, platform launch, risks and market niche. The resulting concepts emphasized a regional startup ecosystem, a research-oriented hybrid marketplace or a global marketplace with stricter entry requirements.

**Discussion:** The concepts combine the tensions in different ways and highlight competing priorities in governance, openness, assurance, scalability and early platform growth.

**Conclusion:** Stakeholders identified recurring design choices for a DTx software-component marketplace. The concepts provide hypotheses for prototyping and evaluation; the study did not test technical feasibility, market demand, regulatory acceptability or effects on development cost or time.

**KEY MESSAGES:** *WHAT IS ALREADY KNOWN ON THIS TOPIC:* DTx development can require substantial software, evidence-generation and regulatory resources. Reuse of software components could reduce duplicated work, but exchange of reusable components in DTx is not well characterized.

*WHAT THIS STUDY ADDS:* Interviews with 24 digital health experts identified 12 design tensions and informed three author-generated marketplace concepts for different contexts.

*HOW THIS STUDY MIGHT AFFECT RESEARCH, PRACTICE OR POLICY:* The findings can guide prototype specifications and policy questions concerning quality labels, software integration, maintenance and responsibility. Effects on cost, development time, adoption and patient access require empirical evaluation.

## INTRODUCTION

Digital therapeutics (DTx) are software-based therapeutic interventions designed to prevent, manage, or treat diseases and disorders by delivering evidence-based therapeutic programs directly to patients.^1,2^ Unlike general wellness applications, DTx must demonstrate clinical efficacy, positioning them alongside conventional pharmacological treatments.^2,3^ The DTx market is growing rapidly,^4^ with over 360 products commercially available, and 140 prescription DTx approved for patient use as of 2024.^5^

Despite this promise, DTx development remains resource-intensive and fragmented. Regulatory complexity, particularly across European jurisdictions, imposes substantial burdens on developers, especially young companies and startups.^6^ Although modular software design has long been recognized as a strategy for promoting efficiency in software engineering,^7^ the DTx field has yet to develop mechanisms for systematic component exchange. In this study, DTx software components are defined as reusable modules performing specific intervention, measurement, analytics, integration or userinterface functions (Supplementary File 1).

Failure to reuse validated components leads to duplicated engineering effort, wasted capital, and prolonged development timelines.^8^ This fragmentation is compounded by limited cooperation between actors in the DTx ecosystem, including researchers, startups, and established health technology companies.^9^ Reusable components already exist across the DTx ecosystem, and there are calls for strategies to handle configurability and re-use of components to ensure scalability of DTx.^8^ Additionally, some ‘innovation platforms’ seek to address this issue by creating ecosystems of reusable components.^10,11^

We propose a two-sided marketplace to connect component developers (supply-side), with DTx designers (demand-side). While two-sided markets leverage network effects to create value for both market sides simultaneously,^12^ healthcare markets operate under stringent regulatory requirements, quality assurance demands, and data protection obligations, distinguishing them from other consumer technology platforms.^5^ The classic “chicken-and-egg” liquidity problem,^13^ where the initial user base is limited, remains a barrier. Concerns around regulatory compliance, intellectual property protection, and liability allocation also require careful consideration.^14^ Furthermore, any marketplace for health software components must address how its offerings integrate with established interoperability standards^15,16^ such as HL7 FHIR (Fast Healthcare Interoperability Resources)^17^ and regulatory aspects.

To date, systematic examination of the design tensions of a component marketplace from a business model perspective remains underexplored. While existing research has investigated digital health ecosystems^18^ and platform economics in healthcare,^19^ the specific intersection of component reusability and two-sided market design lacks empirical focus. Beyond its business model dimension, such a marketplace is a health informatics challenge: reusable components must expose documented software interfaces, align with interoperability standards such as HL7 FHIR, and preserve evidence and regulatory traceability when integrated into new DTx products that support health and care service provision. Based on the four dimensions of viable business models^20^ we address four research questions (RQs): **(RQ1)** Who are the marketplace users? (**RQ2)** What should the marketplace offer? **(RQ3)** How can it create and deliver value? **(RQ4)** How should it capture value?

## METHODS

### Study design

We conducted a qualitative study using semi-structured expert interviews. The Magic Triangle framework^20^ guided the interview structure and the deductive analysis. Ethical approval was granted by the ETH Zurich Ethics Commission (EK 2025-N-05).

### Participants and recruitment

Participants were recruited using purposive sampling supplemented by snowball recruitment to include expertise in digital health, DTx development, platform economics and health-technology entrepreneurship. Four pilot interviews were used to refine the guide and were not included in the analytic sample (n=24). Participants had no relationship with the interviewer, were recruited by email, and provided written informed consent before the interviews. The sample size was set a priori, balancing the breadth of stakeholder groups against the information power expected from expert participants and therefore recruitment stopped after the planned 24 interviews. Because saturation was not assessed prospectively using a predefined procedure, we do not claim thematic saturation.^21^

Participants represented diverse professional roles, sectors, and countries, with 1-20 years of experience in digital health (See Supplementary file 2 for an overview of participant demographics and a detailed list of participants). The majority (19/24) were based in Switzerland, reflecting the study’s origin.

### Data collection

Interviews were conducted in English by YM (male, MSc student in Management, Technology & Economics, trained in qualitative interviewing and supervised by TK and SM) between October 2025 and January 2026 via 1:1 video conferencing. Each interview lasted approximately 45 to 60 minutes and followed a semi-structured guide organized around the dimensions of business models (the “Magic Triangle”).^20^ Participants were informed about the study purpose, YM’s role, and the research team’s involvement in a university innovation platform for digital health interventions.

Prior to, and during the sessions, participants were shown a short slides-based prototype overview to establish a shared baseline (Supplementary File 3: interview guide and presentation). The opening interview script described a broad DTx marketplace connecting providers with adopters. The subsequent presentation focused on reusable DTx components. Interviews were audio-recorded and transcribed using MAXQDA’s AI-supported transcription tool and completely verified and manually reviewed by the human research team for accuracy and anonymity. Participants reviewed their transcripts prior to analysis; two made minor or substantive clarifications. No interviews were repeated; brief field notes were made after each session.

### Data analysis

We analysed data using hybrid deductive-inductive codebook thematic analysis.^22^ The four Magic Triangle dimensions formed the initial deductive structure, with inductive codes added into a ‘general’ dimension. YM coded all transcripts in MAXQDA and maintained an evolving codebook containing code names, definitions and examples. SM independently applied the developing codebook to the four pilot transcripts and one randomly-selected transcript from the analytic sample. The subset size aligns with established guidelines for establishing consistency in codebook application as proposed by Saunders et al.^23^ YM and SM compared code applications and discussed discrepancies to clarify or revise definitions; no inter-rater reliability statistic was calculated. YM then completed coding of the remaining transcripts. SM and MN reviewed the 76 codes and their organization into 17 parent themes.

To derive the 12 design tensions, SM compared patterns within and across the parent themes and identified issues for which participants described competing priorities or alternative design choices. Candidate tensions were checked against supporting and contrasting extracts, overlapping tensions were merged, and the final set was agreed by SM & MN. The tensions were then used as design axes to construct three contrasting marketplace concepts. These concepts are author-generated analytical archetypes informed by the interviews and platform literature; they are not participant clusters, observed marketplaces or tested business models. Supplementary File 5 provides the coding framework and an audit trail from codes and themes to tensions and concepts.

### Reporting standards

This study is reported in accordance with the Consolidated Criteria for Reporting Qualitative Research (COREQ).^24^ (Supplementary File 4: COREQ checklist).

## RESULTS

The 24 interviews yielded 76 codes, which were organized into 17 parent themes within the four Magic Triangle dimensions and the general category. Comparison within and across these themes yielded 12 design tensions. Figure 1 summarizes the tensions and Supplementary File 5 provides the full coding framework and an audit trail linking codes, parent themes, tensions, and supporting and contrasting extracts.

**Figure 1.**
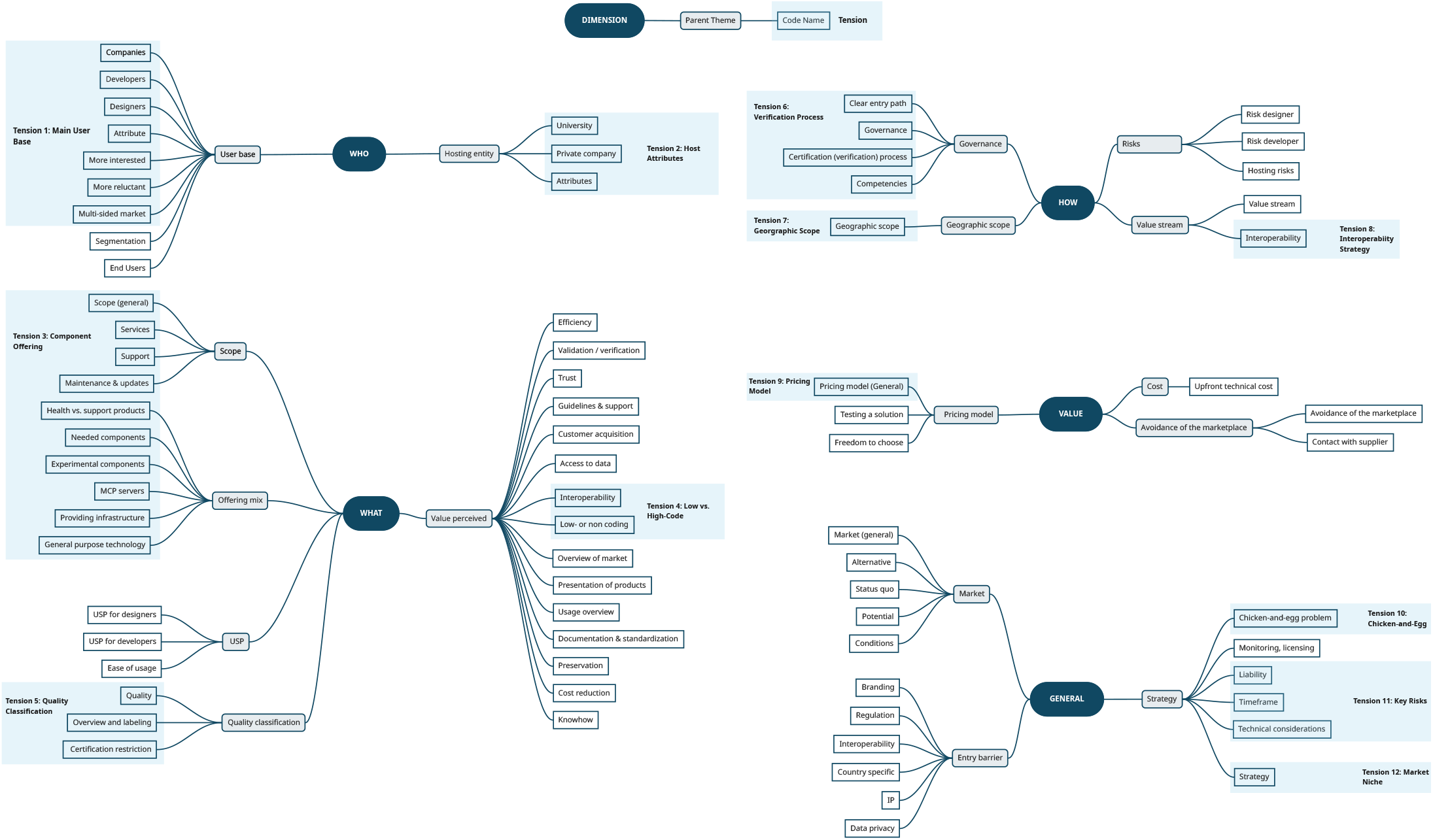
Thematic Map of Dimensions, Parent Themes, Codes and Design Tensions

Synthesizing conceptually related parent themes into higher-level marketplace design trade-offs resulted in twelve design tensions. These tensions served as the analytical bridge between the empirical findings and the three marketplace concepts, and are shown mapped onto the Magic Triangle in Figure 2.

**Figure 2.**
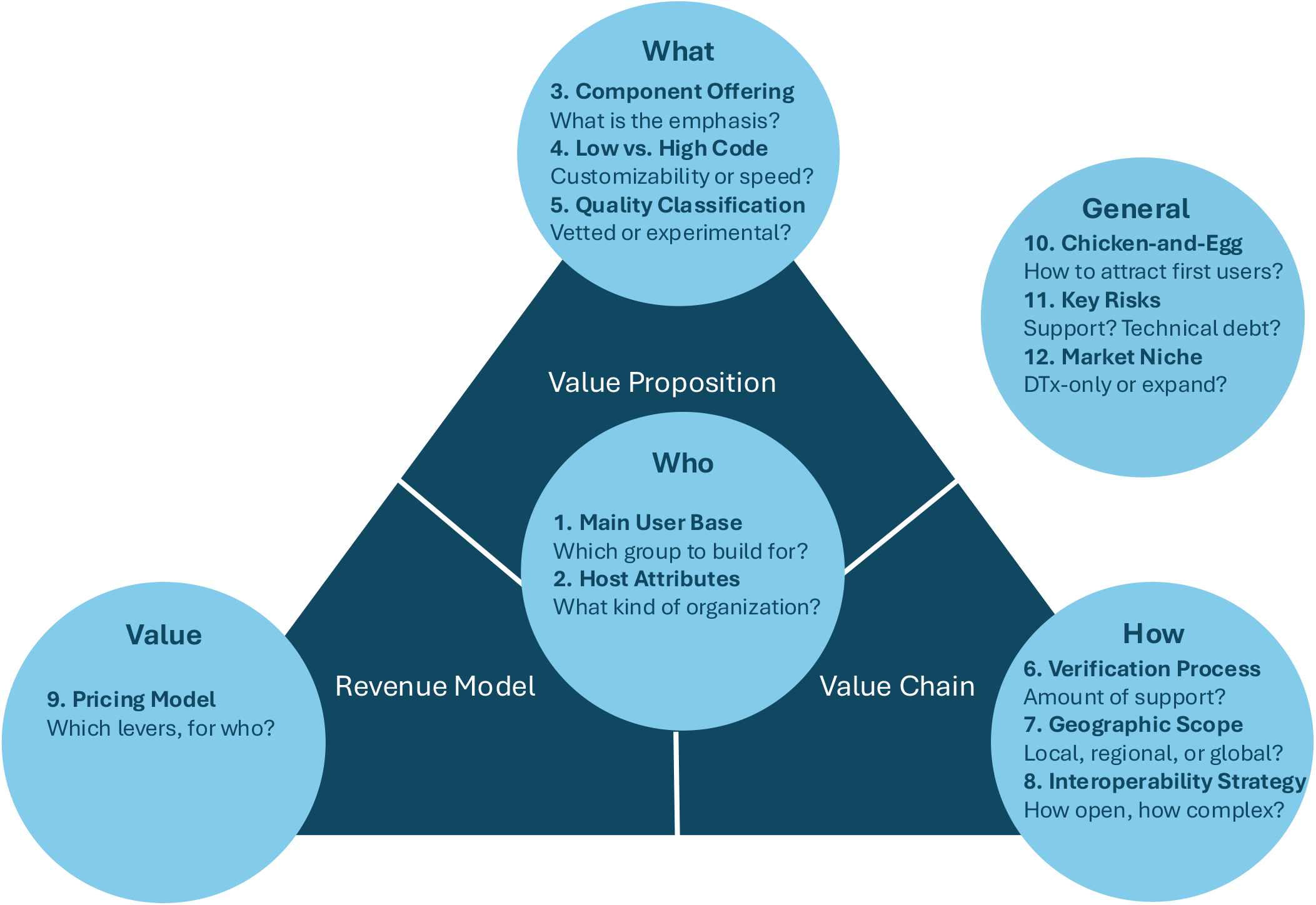
The four dimensions of viable business models (the Magic Triangle)^20^ including the 12 identified key design tensions. Relevant codes which did not fit into the four dimensions were included in the ‘general’ circle.

### Marketplace users

Addressing RQ1, participants identified two primary user bases: component developers (supply side) and DTx designers (demand side), noting that academic groups and SMEs often blur these lines as “prosumers.” Preferences for the hosting entity were equally split between academic institutions, public consortia, and private entities, though host neutrality and credibility were deemed vital for longterm trust:

> *“The technical provider and developer is… important, but its affiliation is secondary, as long as we can trust a long-term relationship*.*”* **(P14, Project Leader)**

### Marketplace offering

Evaluating RQ2, participants discussed a marketplace for reusable software components together with optional support activities, such as documentation, onboarding and maintenance guidance. Software components mentioned included modules implementing validated questionnaires, behavioural interventions, wearable or sensor connections, and user-interface functions. Twelve interviewees indicated that perceived value centered on compressed development timelines and rapid prototyping using pre-validated modules:

> *“You’re much faster on market with it. …You can really use [your development resources for] adding…your magic sauce And if you use well-established components, then of course, this is much easier than if you start from scratch*.*”* **(P9, Product Manager)**

While a low-code architecture was favoured to lower barriers for non-technical designers, experts acknowledged a trade-off with the complexity of use cases:

> “*It gets*… *tricky with low code or no code because, [researchers] want to do something … that hasn’t been done. And the type of customization they need might … require a level of complexity …that is usually overkill, but their specific needs require it*.*”* **(P19, Doctoral Researcher, Biomedical Engineering)**

Participants proposed tiered quality labels distinguishing components with limited testing, components that had passed defined technical or security checks, and components supported by clinical evidence for a specified use. Any label would need to state exactly what had been assessed and should not imply that evidence or regulatory status automatically transfers when the component is integrated into another DTx product:

> “*… It would be good if there’s some validation[*.*] … I’m also fine to use something… if it’s innovative, … that’s not necessarily validated. … I think that’s a bit the point of experimentation, right?*” **(P19, Doctoral Researcher, Biomedical Engineering)**

Beyond component exchange, participants valued the potential for community building, knowledge sharing, and establishing a reference system that could incentivize developers, particularly early-career researchers, to contribute high-quality reusable modules.

### Value creation and delivery

Within the scope of RQ3, participants described two related requirements: a documented software interface that allows components to work together and, where clinical data are exchanged, compatibility with relevant standards such as HL7 FHIR. Participants also described a trade-off between simple developer-facing interfaces at launch and broader standards alignment over time:

> *“Standardization is one of the toughest and most expensive tasks that you can start with… [it] requires highly skilled experts [to] integrate [and it often comes at] the cost of simplicity and interoperability*. … *My recommendation* … *would be to start with a simple, developer-friendly API to lower the entry barrier, but plan a standards adoption in the roadmap. At minimum, use FHIR-compatible data models for clinical data exchange and align with [local patient record] profiles”* **(P24, Managing Partner)**

The verification process emerged as a central governance challenge: participants called for automated malware testing, compatibility verification, and version control mechanisms to ensure that components remain functional as underlying platforms evolve, however, this increasingly puts a burden on developers who submit modules. Onboarding strategies split between open-access models with verification badges and restricted, pre-vetting entry pipelines. Several participants stressed the need for clear documentation standards and transparent onboarding pathways for developers:

> “*At the beginning, you probably can’t be too picky. […] Providing clear guidelines could be a good starting point and would help build trust in your platform*.**” (P24, Managing Partner)**

Finally, while some argued for a localized geographic launch (e.g., the DACH region) to simplify early compliance, most advocated for an immediate international footprint to ensure commercial viability:

> “*Developing products for the small Swiss market is not…really a sustainable business goal. You need, in my opinion, a larger target […]. At least…another EU country like Germany*.” **(P24, Managing Partner)**

## Value capture

Evaluating RQ4, no single revenue model commanded consensus. Subscriptions fit predictable academic budget cycles, while transaction-based fees suited commercial variants but risked platform disintermediation. A freemium strategy was widely suggested to solve initial liquidity problems. One participant with extensive startup experience advised an iterative approach:

> “*I wouldn’t spend too much time trying to find the perfect licensing model, because all the platforms we know change theirs over time [*…*] I’ve also wasted too much time on my own startup trying to perfect a pricing model when we didn’t know anything*.” **(P6, CEO)**

## Inductive codes

Inductively derived codes surfaced topics around attracting a user base, clarifying liability and planning early for scalability.

Participants offered varying opinions about how to beat the “chicken-and-egg” problem, with some advocating a focus on gaining developers first, while others advocated strategic partnerships:

> “*How to make it work? … you would need to first start with the ones that already built… components and connectors and give them an offer…”* **(P7, Chief Product Officer)**
>
> *“[DTx organization] could be another interesting … partner for such a marketplace because they actually created a community around this a partnership [where] you are bringing infrastructure and They have the community and playbooks*.*”* **(P1, CEO)**

Key risks included component support, especially for researcher- or student-developed projects:

> *“[Say] I want to pick those for my study*. .. *until it actually starts,…[it could be] one and a half, two years. And [some of the] companies … God knows what they will be doing in two years*.*”* **(P15, CEO)**

Another risk was that early technical choices could potentially create later obstacles to scaling:

> *“Ignoring standards entirely creates technical debt that becomes exponentially expensive to fix once you need to interoperate with regulated systems*.*”* **(P24, Managing Partner)**

Crucially, experts warned that the standalone DTx market is too narrow, advising an expansion into the broader digital health ecosystem, presenting a trade-off between a niche product with few competitors and customers and a general product with many competitors and customers.

Based on the 12 interview-derived tensions, we constructed three contrasting marketplace concepts. Each concept combines a coherent set of choices rather than representing a participant group or an observed marketplace. Table 1 presents the concepts. The full source mapping for each concept choice is provided in Supplementary File 5.

**Table 1.** Comparative overview of three proposed marketplace concepts for digital therapeutics components derived from the synthesis of interview data and platform economics literature.

| **Tension** | **Regional Concept** | **Hybrid Concept** | **Global Concept** |
| --- | --- | --- | --- |
| **Tension 1:**<br>*Main user base* | Regional startups (developers) and researchers (designers) | Researchers and academic “prosumers” on both sides to seed the platform | International SMEs and scale-ups on both sides |
| **Tension 2:**<br>*Host attributes* | Government-backed consortium to ease regulatory approvals | University-backed consortium, layered on an existing innovation platform | Private company or strategic commercial alliance |
| **Tension 3:** *Component offering* | Moderately open range of component types, optimized for local developers | Broadest range; includes general digital health modules and student projects | Curation restricted to high-quality, verified components |
| **Tension 4:**<br>*Low vs. high code* | Mid-level complexity, balancing compatibility with designer customization | High-code; prioritizes development flexibility over ease of integration | Strict emphasis on low-code, highly-compatible modules |
| **Tension 5:** *Quality classification* | Tiered system; developer verification paired with component self-declaration | Optional host-verification; allows access to experimental modules | Strictest system; mandatory developer and component-level verification |
| **Tension 6:**<br>*Verification process* | Mid-level, well-documented quality assurance; host-supported alignment with local regulatory pathways | Lean, as-needed verification documentation. Low effort for host and developer | Strictest quality assurance, offset by a pre-vetting “enablement platform” for onboarding. |
| **Tension 7:** *Geographic scope* | Tight regional (e.g., DACH: Germany, Austria, Switzerland) | Broader region (e.g. Europe) | Global |
| **Tension 8:** *Interoperability* | Tailored to local standards; higher developer effort but creates platform lock-in | Simple APIs; clinical data standards are optional at launch | Standard APIs plus mandatory compliance with international clinical data standards |
| **Tension 9:** *Pricing model* | Free screening & verification for developers; transaction-based revenue sharing and premium visibility fees | Institutional subscriptions plus transaction-based revenue sharing | Transaction-based revenue sharing; tiered enterprise subscriptions |
| **Tension 10:**<br>*Chicken-and-egg* | Subsidize developers; engage local startups via hackathons and events | Leverage existing innovation platform users; seed with research projects | Strategic alliances with established players; seed with their existing components |
| **Tension 11:**<br>*Key risks* | Small market size; high rework costs if expanding beyond target region | Heavy innovation platform dependence; low-quality components risk user trust and technical debt; Low barrier entry risks marketplace competing with general software hosting platforms | High upfront investment; intense competition from incumbent digital health and EHR giants |
| **Tension 12:**<br>*Market niche* | DTx only with future openness to scale into digital health | DTx and digital health components | A focus on becoming a market leader in DTx components globally. Eventual expansion into broader digital health market |
| **Summary** | Measured, regional approach: optimizes business value to startups, delivering faster local regulatory approval. Compromises market size for a captive user base | Academic flexibility: optimizes for research productivity and customization. Reduces host platform costs but risks high technical debt and variable component quality | Global standardization: optimizes for designer ease of use and interoperability. Requires high upfront investment to manage strict global developer standards |

## DISCUSSION

This qualitative study identified design tensions in a proposed marketplace for reusable DTx software components and used them to construct three analytical marketplace concepts. Its contribution is a stakeholder-informed design space for subsequent prototyping and evaluation.

### Principal findings

Our findings reveal that a DTx component marketplace could add value and time savings to DTx development but faces significant design trade-offs. The most fundamental tension lies between openness and quality control: an open marketplace accelerates growth but risks undermining trust through low-quality components, while stringent verification creates barriers to entry that exacerbate the chicken-and-egg problem. The three proposed marketplace concepts (Table 1) illustrate alternative ways of balancing these trade-offs. They should be interpreted as stakeholder-informed design archetypes derived from the interview data and informed by platform economics literature, rather than as empirically validated marketplace models. Future implementations and real-world evaluations are needed to determine which design strategy is most effective.

### Comparison with existing literature

We extend digital health ecosystem literature^18,19^ by applying two-sided market theory to DTx component reuse. Interoperability requirements emphasized by our participants echo broader calls for health information exchange standards.^15,16^ Standards such as HL7 FHIR can support clinical data exchange, while reusable software components additionally require documented interfaces, dependencies, supported environments, versioning and maintenance responsibilities.

Recent stakeholder research on DTx economic value drivers^25^ corroborates our finding that technical quality assurance, regulatory compliance and clinical validation are considered primary determinants of marketplace viability. By demonstrating how different governance models address identical market needs, these concepts contribute directly to platform design literature. While platform economics predicts “winner-takes-all” consolidation,^26^ our models suggest niche healthcare markets might sustain coexisting, specialized archetypes, given an appropriate geographic scope. This differentiation mirrors the broader observation that successful marketplace strategies must match the maturity and structure of their target ecosystems^27^.

### Implications for practice and policy

For practice, this study addresses a core health informatics challenge: eliminating redundant engineering to accelerate DTx development through component reuse. The concepts provide structured options for prototype design rather than implementation-ready blueprints and should be tested with component developers and DTx designers before decisions are made about governance, quality checks, software interfaces, geographic scope or pricing. For policymakers, they highlight how public-private consortia can promote regional software reuse amidst fragmented digital health settings. The emphasis on standardization (to varying degrees) across all three concepts suggests that policy efforts to promote interoperability standards in digital health could catalyse marketplace development. Furthermore, the regulatory feasibility of reusing certified medical software components across products represents an important area for policy clarification that could lower barriers to marketplace adoption.

### Limitations

This study has several limitations. The sample was predominantly Swiss-based (19/24 participants), many with (in)direct affiliation with a local digital health centre and familiarity with its innovation platform for development and testing of DTx using reusable components,^10^ which may introduce bias toward the marketplace concept. Second, purposive and snowball sampling limit generalizability. Third, one researcher coded all transcripts; although a second researcher independently applied the codebook to five transcripts to establish consistency, no inter-rater reliability statistic was calculated, so the coding may partly reflect the primary coder’s perspective (mitigated by the audit trail in Supplementary File 5). Qualitative data captures strategic intent rather than actual market performance, and rapid advancements in AI development tools may alter the future utility of static software component marketplaces. Finally, the concepts were not generated as participant clusters and or tested for technical feasibility, regulatory acceptability, willingness to participate, pricing, market size or effects on development cost and time.

## CONCLUSIONS

Stakeholders identified 12 design tensions in a proposed marketplace for reusable DTx software components. We used these tensions to construct three contrasting marketplace concepts for different contexts, providing a structured basis for prototype development and comparative evaluation. The interviews do not establish technical feasibility, commercial viability, regulatory acceptability or efficiency gains. Future work should test the concepts with a broader international sample, explicitly consider platform dynamics, evaluate actual software-component integration and maintenance, and clarify evidence and regulatory responsibilities.

## Acknowledgements

We thank all interview participants for their time and insights. Preliminary findings will be presented as a late-breaking abstract at the ISRII 2026 Scientific Meeting, Newcastle, Australia.

## Contributors

TK conceived and designed the study. YM conducted the interviews, developed the codebook iteratively with SM, and performed the primary segment-level thematic analysis in MAXQDA under the supervision of SM and TK. SM led the adaptation of the findings into the journal manuscript format, with conceptual framing contributions from MN and TK. All authors contributed to interpretation of findings, critically reviewed the manuscript, and approved the final version. TK is the guarantor. Beyond the AI-supported transcription described in the Methods, generative artificial intelligence (Gemini) was used for language editing in the preparation of this manuscript.

## Patient consent for publication

Patients or members of the public were not involved in the design, conduct, or reporting of this research.

## Ethics approval

This study involves human participants and was approved by the ETH Zurich Ethics Commission (EK 2025-N-05). Participants gave informed consent to participate before taking part.

## Provenance and peer review

Not commissioned; externally peer reviewed.

## Data availability statement

Anonymised interview transcripts are available upon reasonable request to the corresponding author, subject to the ethical approval conditions of the ETH Zurich Ethics Commission.

## Supplemental material

The following supplementary materials are available online: (1) Short typology of DTx Components; (2) Participant overview and demographics; (3) Interview guide and slides presented to participants; COREQ checklist; (5) Coding framework and an audit trail from codes and themes to tensions and concepts.

## REFERENCES

1. Jacobson NC., Kowatsch T, Marsch LA. Digital therapeutics for mental health and addiction : the state of the science and vision for the future. Published online 2023:254. doi:10.1016/C2020-0-02801-X

2. Fürstenau D, Gersch M, Schreiter S. Digital Therapeutics (DTx). Business & Information Systems Engineering 2023 65:3. 2023;65(3):349–360. doi:10.1007/S12599-023-00804-Z

3. Huh KY, Oh J, Lee S, Yu KS. Clinical Evaluation of Digital Therapeutics: Present and Future. Healthc Inform Res. 2022;28(3):188–197. doi:10.4258/hir.2022.28.3.188

4. Grand View Research. Digital Therapeutics Market (2025-2030). 2025. Accessed July 17, 2026. https://www.grandviewresearch.com/industry-analysis/digital-therapeutics-market

5. Armeni P, Polat I, De Rossi LM, Diaferia L, Meregalli S, Gatti A. Exploring the potential of digital therapeutics: An assessment of progress and promise. Digit Health. 2024;10. doi:10.1177/20552076241277441

6. Draghi M. The future of European competitiveness: A competitiveness strategy for Europe. http://www.commission.europa.eu. Published online 2024. doi:10.2872/1823372

7. Baldwin CY., Clark KB. Design Rules. Volume 1, The Power of Modularity. MIT Press; 2000. doi:10.7551/mitpress/2366.001.0001

8. May R, Glauser R, Denecke K. Identifying Reusable Core Assets of Digital Health Apps. Stud Health Technol Inform. 2024;316:78–82. doi:10.3233/SHTI240350

9. Carrera A, Zoccarato F, Mazzeo M, et al. What drives patients’ acceptance of Digital Therapeutics? Establishing a new framework to measure the interplay between rational and institutional factors. BMC Health Serv Res. 2023;23(1). doi:10.1186/s12913-023-09090-7

10. Kowatsch T, Fleisch E. PRECIOUS: A Design and Trial Service for Precision Digital Therapeutics. Digital Health Zurich (DiZH) Webinar.November 12, 2025. Accessed April 16, 2026. https://www.digitalhealthzurich.ch/sites/default/files/dokumente/251211-PRECIOUS-DHZ-Webinar-Kowatsch.pdf

11. Schmiedmayer P, Bauer A, Zagar P, Ravi V, Aalami O. Stanford Spezi. Accessed July 17, 2026. https://github.com/StanfordSpezi

12. Eisenmann T, Parker GG, Van Alstyne MW. Strategies for Two-Sided Markets. Harv Bus Rev. Published online October 2006. Accessed April 16, 2026. https://hbr.org/2006/10/strategies-for-two-sided-markets

13. Caillaud B, Jullien B. Chicken & egg: competition among intermediation service providers. RAND Journal of Economics. 2003;34(2):309–328. doi:10.2307/1593720

14. Hagiu A. Strategic Decisions for Multisided Platforms. MIT Sloan Manag Rev. 2013;(Winter 2014). Accessed April 16, 2026. https://sloanreview.mit.edu/article/strategic-decisions-for-multisided-platforms/

15. Lehne M, Sass J, Essenwanger A, Schepers J, Thun S. Why digital medicine depends on interoperability. NPJ Digit Med. 2019;2(1):79. doi:10.1038/s41746-019-0158-1

16. Adegoke K, Adegoke A, Dawodu D, et al. Interoperability as a Catalyst for Digital Health and Therapeutics: A Scoping Review of Emerging Technologies and Standards (2015–2025). Int J Environ Res Public Health. 2025;22(10). doi:10.3390/ijerph22101535

17. Health Level Seven International (HL7). FHIR - Fast Healthcare Interoperability Resources Standard. Accessed July 17, 2026. https://www.hl7.org/

18. Huettemann R, Sevov B, Meister S, Fehring L. How to establish digital health ecosystems from the perspective of health service-organizations: A taxonomy developed based on expert interviews conducted as modified Delphi approach. Digit Health. 2024;10. doi:10.1177/20552076241271890

19. Hermes S, Riasanow T, Clemons EK, Böhm M, Krcmar H. The digital transformation of the healthcare industry: exploring the rise of emerging platform ecosystems and their influence on the role of patients. Business Research. 2020;13(3):1033–1069. doi:10.1007/s40685-020-00125-x

20. Gassmann O, Frankenberger K, Choudury M. The Business Model NavigatorlJ: The Strategies behind the Most Successful Companies. Pearson; 2020.

21. Braun V, Clarke V. To saturate or not to saturate? Questioning data saturation as a useful concept for thematic analysis and sample-size rationales. Qual Res Sport Exerc Health. Routledge. 2021;13(2):201–216. doi:10.1080/2159676X.2019.1704846

22. Braun V, Clarke V. Using thematic analysis in psychology. Qual Res Psychol. 2006;3(2):77–101. doi:10.1191/1478088706qp063oa

23. Saunders CH, Sierpe A, Von Plessen C, et al. Practical thematic analysis: a guide for multidisciplinary health services research teams engaging in qualitative analysis. BMJ. Published online 2023. doi:10.1136/bmj-2022-074256

24. Tong A, Sainsbury P, Craig J. Consolidated criteria for reporting qualitative research (COREQ): a 32-item checklist for interviews and focus groups. International Journal for Quality in Health Care. 2007;19(6):349–357. doi:10.1093/intqhc/mzm042

25. Sapanel Y, Cloutier LM, Tremblay G, et al. A group concept mapping study of stakeholder perspectives on digital therapeutics economic value drivers. NPJ Digit Med. 2025;8(1). doi:10.1038/s41746-025-01600-7

26. Belleflamme P, Peitz M. The Economics of PlatformslJ: Concepts and Strategy. Cambridge University Press; 2021. doi:10.1017/9781108696913

27. Adner R. Match Your Innovation Strategy to Your Innovation Ecosystem. Harv Bus Rev. 2006;4(84):98–107.

