## Supplemental Table 1 for "Design tensions in a two-sided marketplace for reusable digital therapeutics software components: a qualitative interview study"

File 1. A compact typology of reusable software DTx components

**Table 1.** A compact typology of DTx components which could be offered on a two-sided marketplace. Note: This author-developed typology is illustrative. Actual technical, evidence and regulatory requirements depend on the component's intended use, technical role and the DTx product into which it is integrated.

| **Component**  **type** | **Examples** | **Main marketplace challenge** | **Function** | **Integration complexity** | **Regulatory and evidence burden** |
| --- | --- | --- | --- | --- | --- |
| **Intervention components** | Breathing training, motivational interviewing logic | Evidence for the intended use, intellectual property and maintenance | Delivery of therapeutic content | Low | High |
| **Measurement/ sensing components** | Questionnaire modules, Patient-reported Outcome Measures (PROM) collection modules, passive-sensing connectors | Validity, consent, data quality and device compatibility | Collect patient-reported or sensor data | Medium | High |
| **Analytics/ AI components** | Time-series analysis, risk-prediction models, dashboard modules | Validation, bias, transparency and monitoring | Preprocess and analyze data including classification and prediction | Medium | High |
| **User-interface modules** | Navigation, data-entry forms, notifications and visualizations | Usability, accessibility and localization | Enable user interaction and presentation | High | High |
| **Integration components** | EHR connectors, identity and authentication modules, FHIR adapters or API clients | Software interfaces, security, versioning and maintenance | Connect a DTx product to external systems and data sources | High | High |
