## Supplemental Table 2 for "Design tensions in a two-sided marketplace for reusable digital therapeutics software components: a qualitative interview study"

File 2. Participant Demographics and Detailed List of Participants

**Table 2.** Characteristics of interview participants (n=24). Experience refers to years of professional engagement in digital health or a related field.

| **Characteristic** | **Total (n = 24)** |
| --- | --- |
| **Gender** |  |
| Male | 14 (58%) |
| Female | 10 (42%) |
| **Role** |  |
| Doctoral Researcher | 6 (25%) |
| CEO | 4 (17%) |
| Professor | 2 (8%) |
| Postdoctoral Researcher | 2 (8%) |
| Head of Research | 2 (8%) |
| Chief Product Officer | 1 (4%) |
| Senior Scientist | 1 (4%) |
| Assistant Professor | 1 (4%) |
| Project Leader | 1 (4%) |
| Digital Strategist | 1 (4%) |
| Head of Sales | 1 (4%) |
| Managing Partner | 1 (4%) |
| Product Manager | 1 (4%) |
| **Sector** |  |
| Academia | 12 (50%) |
| Startup & Startup Support | 6 (25%) |
| Industry | 4 (17%) |
| Government | 2 (8%) |
| **Country (of residence)** |  |
| Switzerland | 19 (79%) |
| Germany | 2 (8%) |
| USA | 2 (8%) |
| Singapore | 1 (4%) |
| **Years of Experience** |  |
| 1-5 | 8 (33%) |
| 6-10 | 6 (25%) |
| 11-15 | 3 (13%) |
| 16-20 | 6 (25%) |
| Not Reported | 1 (4%) |

**Table 3.** Detailed list of participants including job title, professional sector, country of residence, and years of experience

| **ID** | **Role** | **Sector** | **Country** | **Experience (years)** |
| --- | --- | --- | --- | --- |
| P1 | CEO | Startup | Switzerland | 8 |
| P2 | Postdoctoral researcher | Academia | Switzerland | 2 |
| P3 | Professor | Academia | Switzerland | 5-7 |
| P4 | Doctoral Researcher, Food Science & Biostatistics | Academia | Switzerland | 2 |
| P5 | Doctoral Researcher, Biomedical Engineering & Computational Neuroscience | Academia | Switzerland | 1 |
| P6 | CEO | Startup support | Switzerland | 12 |
| P7 | Chief Product Officer | Industry | Germany / Switzerland | 6 |
| P8 | Head of Research | Government | Switzerland | 20 |
| P9 | Product Manager | Industry | Switzerland | 1-5 |
| P10 | Head of Research | Academia | Germany | 17 |
| P11 | Senior Scientist | Academia | Singapore / Switzerland | 11-12 |
| P12 | Professor | Government | Switzerland | 18 |
| P13 | Assistant Professor | Academia | USA | 9-10 |
| P14 | Project Leader | Industry | Switzerland | Not Reported |
| P15 | CEO | Startup | Switzerland | 6-10 |
| P16 | Doctoral Researcher, Biomedical Engineering, | Academia | Switzerland | 3 |
| P17 | Digital Strategist | Startup | Switzerland | 16 |
| P18 | Postdoctoral researcher | Academia | USA | 7 |
| P19 | Doctoral Researcher, Biomedical Engineering, Neuroscience | Academia | Switzerland | 1 |
| P20 | Doctoral Researcher, Biomedical Engineering, Computer Science | Academia | Switzerland | 4 |
| P21 | Doctoral Researcher, Informatics & Information Systems | Academia | Switzerland | 2 |
| P22 | CEO | Startup support | Switzerland | 16-17 |
| P23 | Head of Sales | Industry | Switzerland | 15-16 |
| P24 | Managing Partner | Startup | Switzerland | 15 |
