## Supplemental File 3 for "Design tensions in a two-sided marketplace for reusable digital therapeutics software components: a qualitative interview study"

File 3. Interview Guide and Presentation

Interviews were semi-structured and therefore some interviews differed from the guide presented below.

Interview Guide

Opening

“Thank you for taking the time to participate in this research. We are investigating the viability of a two-sided digital therapeutics (DTx) marketplace. Before we begin, I would like to explain the concept:

A DTx marketplace is a platform ecosystem that connects DTx developers/providers with institutional hosts (hospitals, insurance companies, corporate wellness programs) who wish to integrate digital therapeutics into their offerings. The marketplace facilitates value creation, coordination, and standardization across the DTx supply chain.

Based on your expertise and experience in [domain], I would like to explore your perspectives on the feasibility, requirements, and challenges of such a platform. This interview will take approximately 45-60 minutes.”

Section 1: WHO (Target Users and Ecosystem Actors)

Questions about the key actors and their roles in a DTx marketplace:

- Who should be the target customers/buyers in a DTx marketplace? What types of organizations would benefit most?
- Who should be potential hosts or intermediaries? (e.g., insurers, hospital networks, corporate wellness providers)
- How would you address the 'chicken-and-egg' problem of attracting both supply (DTx providers) and demand (hosts) simultaneously?
- Should the marketplace operate at a regional (DACH), European, or global level initially?

Section 2: WHAT (Products, Services, and Scope)

Questions about the products and services the marketplace should offer:

- What types of DTx should the marketplace include? (e.g., prescription DTx, consumer DTx, specific therapeutic areas)
- Should the marketplace include only complete DTx products or also components/modules?
- What value would a marketplace provide to DTx developers that they cannot achieve independently?
- What are the critical features or services that would make the marketplace attractive to both sides?

Section 3: HOW (Value Creation and Operations)

Questions about operational mechanisms and value creation:

- What competencies and capabilities must the marketplace operator possess?
- What barriers to entry exist for DTx providers? How could a marketplace reduce them?
- How should DTx quality, efficacy, and safety be verified? What role should the marketplace play?
- What standardization or interoperability requirements are essential? (e.g., data formats, APIs, integration protocols)
- Should the marketplace provide integration services, technical support, or regulatory guidance?

Section 4: VALUE (Revenue Models and Sustainability)

Questions about financial viability and value capturing:

- What revenue models would be viable for a DTx marketplace? (e.g., transaction fees, subscription, freemium)
- What are the major cost drivers for operating a DTx marketplace?
- Would you expect the marketplace to be profitable? If so, over what timeline?
- What factors would be critical to financial sustainability in the long term?

Section 5: General

Additional strategic and contextual questions:

- What competitive threats or alternative models might undermine a centralized DTx marketplace?
- What legal or regulatory hurdles would the marketplace need to navigate?
- What would be the key success factors or critical mass needed to launch successfully?
- What growth strategies would be most effective for scaling the marketplace?
- What are the major risks to the marketplace model, and how might they be mitigated?

Closing

Are there any other aspects of a DTx marketplace that we have not discussed and that you believe would be important to consider? Would you like to add anything to our discussion?

Interview Presentation

The concept of the two-sided marketplace was explained and presented with the help of slides and/or a prototype of a possible website design:

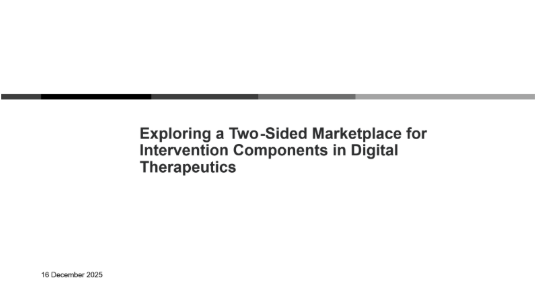

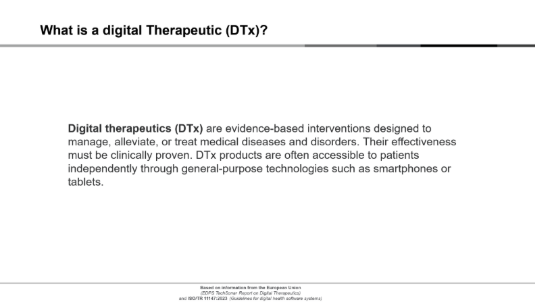

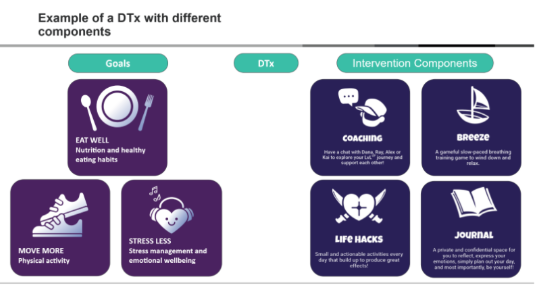

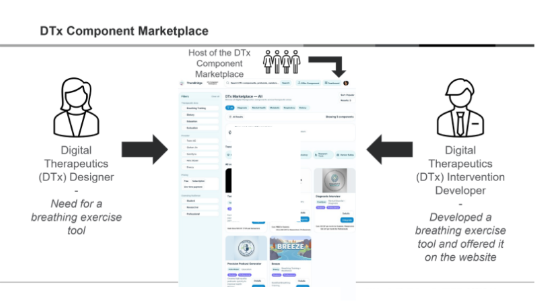

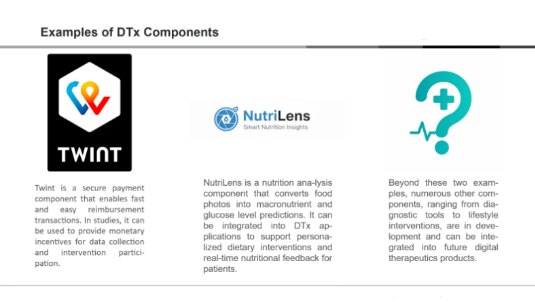
