## Supplemental Table 5 for "Design tensions in a two-sided marketplace for reusable digital therapeutics software components: a qualitative interview study"

File 5. Extended Coding Framework

**Table 4.** Full coding framework and audit trail linking codes, parent themes, tensions, and supporting and contrasting extracts. Themes were subsequently synthesized into the twelve marketplace design tensions presented in Figure 2. These design tensions formed the basis for developing the three stakeholder-informed marketplace concepts described in Table 1 in the main text.

Note: The codes Services and Support refer to marketplace-operator activities and are not treated as DTx component types.

| **Dimension** | **Theme** | **Definition** | **Codes** | **Tension** | **Example Quotes (Anonymised)** |
| --- | --- | --- | --- | --- | --- |
| Who | 1. User base | References to and attributes of potential users of the marketplace | 1. Companies 2. Developers 3. Designers 4. Attribute 5. More interested 6. More reluctant 7. Multi-sided market 8. Segmentation 9. End users | 1. Main user base | “You should start something like the [local digital health center]... or Swiss startups. ... usually the startups ... they're very open to participate in something like that.” (P23, Head of Sales)  “There would probably, a way to go through all the innovation centers in Switzerland ... and also with universities, not only talking about [large ones] but also about smaller ones.” (P22, CEO)  “Smaller scale companies that don't have, the resources to develop such things in-house could potentially leverage on that.” (P11, Senior Scientist) |
|  | 2. Hosting entity | Potential organizations that could host, operate or govern the marketplace, and the attributes participants considered important. | 10. University 11. Private company 12. Attributes | 2. Host attributes | “I would of course prefer as a university institute a partner that it stays in university” (P12, Scientific Director)  “A company like a startup, an AG or something that actually, you know, has a goal to drive profit and keep going. It could be attached to a lab. That's fine. But you need a commercial entity to ensure that this survives on the long term.” (P17, Digital Strategist)  "I'm not sure if it should be a startup or more a governmental organization behind such as [government medical office], some some body that is established, and has yet enough credibility.” (P22, CEO) |
| What | 3. Scope | References to the range of reusable software components listed on the marketplace and to optional support activities provided by the marketplace operator, such as documentation, onboarding, verification or maintenance guidance. | 13. Scope (general) 14. Services 15. Support 16. Maintenance and updates | 3. Component offering | “You could still have a model where they where there are more academic projects or, I don't know, beta projects that could be one section of the marketplace even.” (P1, CEO)  "The easier option for the user would be if everything is in the marketplace, so that you have everything like maintenance and support” (P2, Postdoctoral Researcher)  “You can argue we don't want to overload this platform with these regulation things, we want to focus on the on the functions and these other things are not in the scope of the platform, which is completely fine. But if …you want to support start-ups which don't have a big background, it would be [good to have verified components] to help them to survive this regulatory jungle.” (P10, Head of Research) |
|  | 4. Offering mix | Specific component types needed in the marketplace | 17. Health vs. support products 18. Needed components 19. Experimental components 20. MCP servers 21. Providing infrastructure 22. General purpose technology |  | “A substance use diary or behavioral addiction behaviors, diary so to say....and there should be, of course, AI powered chatbot ... end to end protected video calls or ...internal messaging system that is protected. And so these are, well, I would say the four most important components for me that should be there.” (P12, Scientific Director) |
|  | 5. Value perceived | Value proposition from participants' perspective | 23. Efficiency 24. Validation / verification 25. Trust 26. Guidelines and support 27. Customer acquisition 28. Access to data 29. Interoperability 30. Low-or non-coding 31. Overview of market 32. Presentation of product 33. Usage overview 34. Documentation and standardization 35. Preservation 36. Cost reduction 37. Knowhow | 4. Low vs. high-code | “You want to reduce friction as much as possible, especially because a lot of the people who would be developing these therapeutics and pushing it and even like integrating or downloading in their apps, are not going to be technical people. They would be psychologists. They would be behavioral scientists who have very little technical understanding of how the source code integration would work.” (P13, Assistant Professor)  “It gets a bit more tricky with low code or no code because, [researchers] want to do something … that hasn't been done. And the type of customization they need might … require a level of complexity …that is usually overkill, but their specific needs require it.” (P19, Doctoral Researcher, Biomedical Engineering) |
|  | 6. USP | Unique selling propositions attractive to DTx designers and developers | 38. USP for designers 39. USP for developers 40. Ease of usage |  | “If you use well-established components, then of course, this is much easier than if you start from scratch.” (P9, Product Manager) |
|  | 7. Quality classification | Need for clear labeling, categorization, and overview of components | 41. Quality 42. Overview and labeling 43. Certification restriction | 5. Quality classification | “You could basically introduce categories where...if a component is produced by an ISO certified company, then they can mark it as such.... which gives them some sort of quality assurance... Then you could also introduce user ratings. " (P13, Assistant Professor)  “[If you] go lean for the developers, definitely it's an easy place to launch their products. And, you know, sort of I mean, if they could do that on GitHub and whatnot as well and call it like open source and like open access to everyone. But in terms of if there is a dedicated marketplace for that, to browse and look for things, I think it can be very promising for the developers.” (P16, Doctoral Researcher, Biomedical Engineering)  “If you say okay, you only allow medical products that have ISO certification or [local certified body] registered, then it would be fine. It would be fully trustworthy and then maybe have a higher level, stacked on top where you can put together the modules that are already certified. Then maybe the certification process for the whole thing again is very, very easy. So that would be a trustworthy platform.” (P3, Professor) |
| How | 8. Governance | Governance structures and policies for marketplace operation | 44. Clear entry path 45. Governance 46. Certification (verification) process 47. Competencies | 6. Verification process | “You can set up different services around it, regulatory legal services, but also, the whole yeah, certification process, which is quite a pain for all the healthcare startups. (..) For most of them, that could be a nice add on on the platform. Yes. Why not? But then again, shouldn't it be like different people from Swiss medtech, from BAG, from eHealth Swiss, from other other organizations that are that run the platform or are and stand behind them personally, but also with their organizations.” (P22, CEO)  “Make it so like such a low threshold for them to put up their tools that they may already have … like to attract people who already have maybe open source tools and just give them a thing where you say, well, you know, you can just repackage your open source tool that you already have” (P19, Doctoral Researcher, Biomedical Engineering) |
|  | 9. Geographic scope | Preferred geographical scope for the marketplace (local, DACH, EU, global) | 48. Geographic scope | 7. Geographic scope | “We know the Swiss, path to get approvals. So it would be great to have sort of, feedback from the regulators about the marketplace. Then then you probably need to do some interviews with the [local certified body] and these regulator side because, because then it could be very … for companies which are just focused on the Swiss market. (P1, CEO)  “Developing products for the small Swiss market is not, is not  really a sustainable business goal. You need, in my opinion, a larger target for that. Aiming at least to another EU country like Germany.” (P24, Managing Partner)  “Well, what you shouldn't and must not do is to only focus on Swiss solutions. So the question is how much do I open this this platform? Is it only for the DACH region? Is it for Europe? I'd rather leave it open for anyone. If the regulatory and quality checks are are nicely done then nothing, in my honest opinion, nothing speaks against the full opening.” (P22, CEO) |
|  | 10. Risks | Risks associated with using, hosting or operating the marketplace | 49. Risks designer 50. Risks developer 51. Hosting risks |  | “If I have the funding as a designer and I want to have something designed, I would like to give the task to somebody who then takes responsibility, for the for the end product. And with this platform, you know, I'm the person that takes responsibility at the end, but then I have limited visibility.” (P15, CEO) |
|  | 11. Value stream | Value creation and delivery mechanisms of the marketplace | 52. Value stream 53. Interoperability | 8. Interoperability strategy | “Because of the re-development costs and their complexity, those  software products achieve high degree of integration in the existing products. There are also several reasons why reusability and integration of third-party software is not always the best choice. One of the main barrier is that most of the software still in use today is installed on prem increasing the operational complexity (network access, distributed updates, customization, local integration, etc). ..IHE and FHIR, electronic patient record is referencing many of these integration profiles (not proper standards) which are designed to be flexible and extensible, but ofen these come to the cost of simplicity and interoperability guarantees. I would not care about this too much. In my opinion, the market will find the right level of adoption of the standards. What you can still do, if you follow the open API path, is to leverage the information that you will accumulate to promote defector standards. Reducing the barrier to integrate with regional or national systems and generating tangible value for your platform members when targeting a specific markets or ecosystems. ” (P24, Managing Partner)  “You want to reduce friction as much as possible, especially because a lot of the people who would be developing these therapeutics... are not going to be technical people. ... They'll have to either work with a technical team, or hire a software developer who can integrate it into the platform. ... So ... you support only for your platform and it works really well for your platform. But the limitation is that people have to be on your platform (P13, Assistant Professor)  “There was an exchange with some experts and initiatives from Sweden and one of their main platforms that... enabled to faster digitize the healthcare industry was a national platform where all the solution providers had to adopt to the standards of the platform and not bringing in their own. Some of them were quite happy about that because, yes, standardization is the is the basic... for a fast digitization. Others were not too happy, but in the end they ... achieved the goal. And I think 95% of the, solutions that applied for the platform, they all, they actually all committed to, to the standards and like that day they could really standardize also the solution. So that's that's the way to go for me as well. (P22, CEO) |
| Value | 12. Pricing models | Mechanisms for capturing value and generating revenue | 54. Pricing model (general) 55. Testing a solution 56. Freedom to choose | 9. Pricing model | “People hate subscription[s], but it might be the most scalable” (P16, Doctoral Researcher, Biomedical Engineering)  “The one time payment plus the subscription could also be able to offer both. So for starters, they will not. Nobody will subscribe because they've never worked with you. So. But perhaps you will then move people into subscription. If they say, hey we use that a lot, then of course subscription makes sense. So I would, I would, I would be offering different entry points from the start and at the same time. (..)” (P6, CEO)  “You pay the marketplace a subscription fee to be able to offer your products, but then gain all the money from the product… I think I would prefer [that] option. (P8, Head of Research)  “My recommendations is be flexible build a hypothesis. Say hey this is what we think will work on this platform based on research we've done, based on other platforms and so on, but we're not 100% sure. Hence we're going to try it out for this long and then we're going to swap it. Be open about it. Be. This is our pilot processing model period. And we're free to adjust it. I think this too much and I've also wasted too much time in my own startup perfecting a pricing model and we didn't know anything.” (P6, CEO) |
| Value | 13. Upfront technical costs | Major cost drivers for marketplace operations | 57. Cost |  | "it will cost you right to, to do to do this on a professional level. This will cost money. So you need some sort of model where at least you can pay the cost that that this project will cost." (P6, CEO) |
|  | 14. Avoidance of the marketplace | Reasons stakeholders might avoid using the marketplace | 58. Avoidance of the marketplace 59. Contact with suppliers |  | "[circumvention of the marketplace] would be difficult to avoid... ... Sometimes we also have limited amount of money from the budget wise, just from the, the founders. And then of course, we have to find a good cheap solution. And if it's cheaper, well honestly, maybe we will ask directly....  You should also consider not only to sell it to potential designers, but to whole maybe stakeholder groups or so. ...On a different level...  Interviewer: “You mean like offering the product that you have a subscription as a university and then you have access to all the components? “  "Exactly.” (P12, Scientific Director) |
| General | 15. Strategy | General marketplace launch, growth and governance strategies | 60. Chicken-and-egg problem 61. Monitoring, licensing 62. Liability 63. Timeframe 64. Technical considerations 65. Strategy | 10. Chicken-and-egg  11. Key risks  12. Market niche | Chicken-and-egg:  “Ultimately, I think I think you would need to first start with the ones that already built, like successful components and connectors and give them an offer, like to monetize to find further monetization potential for that.” (P7, Chief Product Officer)  “A possibility is starting with students. Because I think during master thesis the people are like like it's it's barely the case that someone has half a year to focus on a certain project. And in this case, everybody has to do it. So they are not like doing it for the big bucks, but they are doing it for the, for the ECTS and often they are really nice products coming out of it which really can enhance and are innovative. So like this you could like gain a lot of projects faster and they still have like they they do it for the ECTS, so even if they would then get $1 or one francs each someone is using it.” (P21, Doctoral Researcher, Informatics and Information Systems)  Key risks:  (losing trust) “You need to make sure that, like, if I'm a young DiGA developer with a great idea and I'm actually going to invest some money in getting components in to get started, that I can also trust that this stuff is not a scam, that this actually works, that this has no security flaws and so on” (P7, Chief Product Officer)  (competition with general marketplaces) “How does it then compare to the low no-code platforms? Does it offer a difference in in the value or the benefit that is strong enough to attract both sides of the market or not.” (P17, Digital Strategist)  (component support) “[Say] I want to pick those for my study. .. until it actually starts,…[it could be] one and a half, two years. And [some of the] companies … God knows what they will be doing in two years.” (P15, CEO)  (technical debt) “Ignoring standards entirely creates technical debt that becomes exponentially expensive to fix once you need to interoperate with regulated systems.” (P24, Managing Partner)  Market niche:  “Are you trying to achieve a vast portfolio of great plugins, features, whatever you name it, then I wouldn't restrict it. Right. But then this would be a marketplace for basically app development in healthcare. And this would be broader. It would not just be DTx.” (P6, CEO)  “You want to have a marketplace where, you know, there's a big market. I don't know, a lot of people have children and a lot of people have old stuff from their, when their were children were younger. So, you know, they are obviously two parties who can bring together. There you have a marketplace. Here it seems like a very niche solution. So marketplace is maybe too big of a word for that.” (P15, CEO) |
|  | 16. Market | General market context and dynamics | 66. Market (general) 67. Alternative 68. Status quo 69. Potential 70. Conditions |  | “I would urge everybody who's working on that to think really carefully about the markets for this solution and think carefully about, not being in a bubble, because I can see how you can get 20 digital therapeutic people in one room and they will be all excited about it, but the minute there is some poor soul who's responsible for that and who founds a company and tries to monetize that, it will be a harsh awakening for them." (P15, CEO) |
|  | 17. Entry barrier | Reasons why potential users might choose not to enter the marketplace | 71. Branding 72. Regulation 73. Interoperability 74. Country specific 75. IP 76. Data privacy |  | “I would have questions about who's making money from this, and how if my intervention is built on your platform, who has all the data and where does all that data go? And do you have like rights over that data or not?” (P11, Senior Scientist) |
